# Respiratory infection during lithium and valproate medication: a within-individual prospective study of 50,000 patients with bipolar disorder

**DOI:** 10.1101/2020.05.04.20090084

**Authors:** Mikael Landén, Paul Lichtenstein, Henrik Larsson, Jie Song

**Affiliations:** Institute of Neuroscience and Physiology, The Sahlgrenska Academy at Gothenburg University, Gothenburg, Sweden; Department of Medical Epidemiology and Biostatistics, Karolinska Institutet, Stockholm, Sweden; Department of Medical Sciences, Örebro University, Örebro, Sweden

**Keywords:** Respiratory tract infections, Lithium, drug repositioning, viruses, therapeutic use

## Abstract

**Objective:** *In vitro* studies have demonstrated that lithium has antiviral properties, but evidence from human studies is scarce. Lithium is used as a mood stabilizer to treat patients with bipolar disorder. Here, the aim was to investigate the association between lithium use and the risk of respiratory infections in patients with bipolar disorder. To rule out the possibility that a potential association could be due to lithium’s effect on psychiatric symptoms, we also studied the effect of the most common alternative to lithium to prevent mood episodes in bipolar disorder, valproate.

**Method:** We followed 51,509 individuals diagnosed with bipolar disorder in the Swedish Patient register 2005–2013. We applied a with-individual design using stratified Cox regression to estimate the hazard ratios (HRs) of respiratory infections during treated periods compared with untreated periods.

**Results:** During follow-up, 5,760 respiratory infections were documented in the Swedish Patient Register. The incidence rate was 28% lower during lithium treatment (HR 0.73, 95% CI 0.61–0.86) and 35% higher during valproate treatment (HR 1.35, 95% CI 1.06–1.73) compared with periods off treatment.

**Conclusions:** This study provides real-world evidence that lithium protects against respiratory infections and suggests that the repurposing potential of lithium for antiviral effects is worthy of investigation.

## Introduction

The outbreak of the coronavirus SARS-CoV-2, the cause of COVID-19 (Corona virus Disease-2019) respiratory disease, has prompted a search for approved drugs with antiviral properties that might be repurposed to treat covid-19, or shed light on biological mechanisms that might be targeted for antiviral purposes.

Lithium has been suggested to have antiviral properties since the 1980s.^1 2^ *In vitro* studies have shown that lithium inhibit replication of several viruses including but not limited to herpesvirus type 1,^3^ transmissible gastroenteritis virus,^4^ reovirus,^5^ food-and-mouth disease virus,^6^ and avian leukosis virus subgroup J.^7^ Interestingly, *in vitro* studies have also shown lithium to effectively suppress infection with Porcine deltacoronavirus^8^ and other viruses that belong to the Coronaviridae family, albeit usually in concentrations that would be toxic in humans^9^.

There is, however, scarce evidence to support that lithium affects viral diseases in humans. Small studies in the 1990s suggested reduced rates of herpes virus infections with lithium.^10–12^ A retrospective review of 236 patients with affective disorders found that lithium treated patients showed a small reduction of flu-like illness during lithium therapy.^13^

Lithium is the mainstay prophylactic treatment of bipolar disorder.^14^ Here, we leveraged a Swedish cohort of more than 50,000 patients with bipolar disorder followed over eight years to investigate if lithium treatment is associated with decreased risk for respiratory infections. We employed a within-individual design to circumvent the problem of confounding by indication. With each person serving as his or her own matched control, the within-individual design controls for fixed confounders (e.g., baseline severity of the illness, genetic predisposition, childhood environment).^15^ To rule out the possibility that a potential association between lithium and respiratory infection rate is secondary to the effect on psychiatric symptoms, we also calculated the association between valproate medication and respiratory infections. Valproate is a commonly used alternative to lithium to prevent mood episodes in bipolar disorder^16^, recommended by the National Institute for Health and Care Excellence (NICE) when lithium is not suitable.^17^

## Methods

### Subjects

We linked several longitudinal Swedish population-based registers through unique personal identification numbers:^18^ the Total Population Register, the Migration Register, the Cause of Death Register, the Patient Register (captures information on inpatient care since 1973 and outpatient visits to specialists care since 2001), and the Prescribed Drug Register.^19^ We identified 51,509 individuals with bipolar disorder followed from 1 October 2005, or age 15, or date of bipolar disorder diagnosis (if later than 1 October 2005) until emigration, death, or 31 December 2013, whichever occurred first. This study was approved by the Ethics Committee at Karolinska Institutet.

### Bipolar disorder cases

To identify cases with bipolar disorder in the Patient Register, we applied a validated algorithm^20^ that has been used in several previous Swedish register-based studies on bipolar disorder.^21–23^ This algorithm includes bipolar I disorder, bipolar II disorder, bipolar disorder not otherwise specified, or schizoaffective disorder bipolar type.

### Medications

The main exposure was defined as dispensed prescriptions of lithium sulphate (Anatomical Therapeutic Chemical classification code: N05AN01) or sodium valproate/valproic acid (N03AG01) as documented in the prescribed drug register. In Sweden, long-term prescribed medications are dispensed for three-month periods. Similar to previous studies,^22^ we therefore defined a medication period as a sequence of at least two prescriptions, with no more than three months (92 days) between any two consecutive prescriptions. Thus, for both lithium and valproate, individuals were defined as on medication during the time interval between two dispensed prescriptions, unless the dispensed prescriptions occurred more than three months apart. To determine whether an individual was on/off medication initially, the start of follow-up was set to 1 October 2005 because the prescribed drug register started 1 July 2005.

### Outcomes

In Sweden, diagnoses of uncomplicated influenza or other viral respiratory infection treated outside hospital are made in primary care and therefore not recorded in the Patient register. When a viral respiratory infection is severe enough to warrant hospital treatment, it is commonly due to a complicating secondary bacterial infection. Hence, patients with a primary viral respiratory infection are rarely diagnosed as such, but are more likely to be discharged with a diagnosis of bacterial pneumonia. We therefore defined outcome as diagnoses of respiratory infections due to virus or bacteria. Specifically, the outcomes were influenza and other viral pneumonia (ICD-10 codes: J09, J10, J11.0, J11.1, J12), bacterial pneumonia (ICD-10 codes: J13, J14, J15, J16), pneumonia in other diseases (ICD-10 code: J17.1), pneumonia due to unspecified organisms (ICD-10 code: J18), acute bronchitis (ICD-10 code: J20), acute bronchiolitis (ICD-10 code: J21), and unspecified acute lower respiratory infection (ICD-10 code: J22). In an additional posthoc analysis, we limited outcomes to respiratory infection diagnoses specifically caused by viruses (ICD-10 codes: J09, J10, J11.0, J11.1, J12, J17.1, J20.3-7, and J21.0-1). Dates and diagnoses were retrieved from the National Patient Register.

### Statistical analyses

To control for time-invariant covariates, we performed within-individual analyses using stratified Cox regression. This method only obtains information from individuals who ever had a respiratory infection during the follow-up; individuals without any of the outcome diagnoses during follow-up do not provide information. More details are given in a review.^24^

We split the follow-up time into consecutive periods. A new period started after a medication switch (i.e., from off medication to on medication or vice versa, for either lithium or valproate) or a respiratory infection diagnosis. For the latter, we restarted the following period at baseline (i.e., to set the underlying time scale as the time since last diagnosis). Lithium and valproate treatment status were defined as time-varying dichotomous exposures. Age band (15-51, 51-64, 64-75, 75-100 years, grouped by the quartiles of the baseline age distribution among those who had an event during the follow-up) was adjusted for as a time-varying categorical covariate. We estimated the hazard ratios (HRs) and 95% confidence intervals (CIs) for respiratory infections during periods with treatment as compared with periods off treatment. This method have previously been described in detail.^15 25–27^

All analyses were performed with Stata 16.0.^28^

## Results

Among the 51,509 patients with bipolar disorder, a total of 5760 respiratory infections diagnoses occurred in 3389 individuals (6.6%) during 217,214 person-years of follow-up (Table 1). Lithium treatment was more prevalent (41.0%) than valproate treatment (15.9%). About 50% patients were not exposed to lithium or valproate during the study period.

**Table 1.** Characteristics at baseline and during follow-up for patients with bipolar disorder with different medication status in Sweden 2005-2013.

|  | **Lithium<sup>a</sup>** | **Valproate<sup>a</sup>** | **Never treated with lithium / valproate** | **Total** | **Eligible for within-individual analysis<sup>c</sup>** |
| --- | --- | --- | --- | --- | --- |
| **Number of individuals (%)** | 21,126 (41.0) | 8,410 (15.9) | 25,758 (49.7) | 51,509 | 3,285 (6.4) |
| **Sex (% male)** | 8,469 (40.1) | 3,609 (42.9) | 9,001 (34.9) | 19,475 (37.8) | 1,307 (39.8) |
| **Characteristics at baseline** |  |  |  |  |  |
| **Age distribution (N of individuals, %)<sup>b</sup>** |  |  |  |  |  |
| 15–51 | 11,355 (53.7) | 5,115 (60.8) | 16,263 (63.1) | 3,0808 (59.8) | 864 (26.3) |
| 51–64 | 5,513 (26.1) | 1,998 (23.8) | 4,986 (19.4) | 11,494 (22.3) | 899 (27.4) |
| 64–75 | 2,652 (12.6) | 887 (10.5) | 2,498 (9.7) | 5,496 (10.7) | 798 (24.3) |
| 75–100 | 1,606 (7.6) | 410 (4.9) | 2,011 (7.8) | 3,711 (7.2) | 724 (22.0) |
| **Respiratory infection (N of individuals, %)** |  |  |  |  |  |
| At least one respiratory infection event during follow-up | 1,532 (7.3) | 722 (8.6) | 1,463 (5.7) | 3,389 (6.6) | 3,285 (100) |
| **Proportion of time exposed to lithium or valproate medications** | 41.3 | 42.3 | - | 22.4 | 28.6 |

- a. 3,785 individuals were ever treated with both lithium and valproate (defined as at least 3 months treatment) - b. Age band was categorized according to the quartiles of age distribution of individuals who ever have any respiratory infection at any time during follow-up. - c. Includes individuals who ever had a respiratory infection during the follow-up; individuals without any of the outcome diagnoses during follow-up do not provide information.

After excluding individuals without respiratory infections diagnoses and varying covariates (i.e., treatment status and age band) during follow-up, 3285 individuals were eligible for the within-individual analysis. These patients had 5656 respiratory infection diagnoses.

The rate of respiratory infection diagnoses was 27% lower during periods on lithium treatment compared with periods off lithium (HR 0.73, 95% CI 0.61–0.86). By contrast, the rate of respiratory infection diagnoses was higher during valproate treatment (HR 1.35, 95% CI 1.06–1.73).

When we, in a posthoc analysis, limited the diagnoses to respiratory infections explicitly caused by viruses, the number of events dropped by 96% to 199. In this analysis, neither lithium (HR 1.07, 95% CI 0.35–3.22) nor valproate (HR 0.96, 95% CI 0.24–4.37) was significantly associated with the risk of viral respiratory infections.

## Discussion

In a large sample of patients with bipolar disorder, we found that rates of respiratory infection diagnoses were significantly decreased while on lithium medication compared with periods off lithium. By contrast, the rate of respiratory infection diagnoses was increased during medication with the alternative mood stabilizer valproate. Given that we used a within-individual design—and that lithium and valproate both effectively prevent mood episodes in bipolar disorder—the difference is likely to be caused by the medication.

The observed inverse association between lithium and respiratory tract infections aligns with the notion based on *in vitro* studies that lithium has antiviral properties.^1 29^ Our findings are also in line with a small previous study (N=236) that found a reduction of flu-like illness during lithium therapy.^13^ Although lithium is believed to act across several biological pathways and the exact mechanism of actions remains obscure,^30^ it is known to inhibit glycogen synthase kinase 3β (GSK3β).^31^ GSK3β has been suggested to be involved in inducing apoptosis and release of viral particles in dengue virus-2 infection,^32^ and GSK3β suppression is hence one potential mechanism underlying the antiviral effect of lithium.^33^ Lithium has also been found to induce leukocytosis,^34 35^ and attenuate anti-inflammatory effects during chronic use (reviewed by ^2^). Further, lithium may stimulate immunoglobulin production in B-lymphocytes, increase T-lymphocyte proliferation, and affect proinflammatory cytokine production (reviewed by^29^ and^2^).

Valproate treatment, by contrast, *increased* the risk of respiratory infections. It is well known that valproate can cause reversible bone marrow suppression leading to leukopenia and neutropenia.^36^ However, a meta-analysis of infectious diseases reported as adverse events during the double-blind phase of placebo-controlled randomized clinical studies failed to show a significant effect of valproate.^37^

This is by far the largest study testing the hypothesis that lithium protects against respiratory infections. By comparing treated and untreated periods within the same individual, our approach automatically controlled for all time-stationary confounders and thus reduces the likelihood of confounding by indication. A limitation is that we not only included diagnoses of respiratory infections due to viruses but also other infectious agents. This was because we relied on hospital diagnoses, and patients with viral respiratory infections are usually admitted to hospital because of secondary bacterial infections. Since patients are rarely tested for, e.g., influenza virus, the discharge diagnosis tends to be bacterial pneumonia or other bacterial respiratory infection despite a primary viral infection. This is echoed by the dramatic decrease of number of events (97%) when we limited the analysis to explicit viral respiratory infections in a posthoc analysis. This radically lowered statistical power and the posthoc analysis did not yield any significant results. Hence, it cannot be excluded that lithium protects against other causes of respiratory infections than viruses. Limitations finally include the lack of information regarding adherence, but this would if anything be a conservative bias.

In conclusion, we provide real-world evidence that lithium treatment reduces the rate respiratory infections. Taken together with ample previous literature on antiviral effects of lithium, the repurposing potential of lithium for antiviral effects is worth pursuing.

## Data Availability

The authors had full and ongoing access to the original data presented and analysed in this study. Due to Swedish legal restrictions, register data cannot be shared.

